# Social Media Study of Public Opinions on Potential COVID-19 Vaccines: Informing Dissent, Disparities, and Dissemination

**DOI:** 10.1101/2020.12.12.20248070

**Authors:** Hanjia Lyu, Junda Wang, Wei Wu, Viet Duong, Xiyang Zhang, Timothy D. Dye, Jiebo Luo

## Abstract

The current development of vaccines for SARS-CoV-2 is unprecedented. Little is known, however, about the nuanced public opinions on the coming vaccines. We adopt a human-guided machine learning framework (using more than 40,000 rigorously selected tweets from more than 20,000 distinct Twitter users) to capture public opinions on the potential vaccines for SARS-CoV-2, classifying them into three groups: pro-vaccine, vaccine-hesitant, and anti-vaccine. We aggregate opinions at the state and country levels, and find that the major changes in the percentages of different opinion groups roughly correspond to the major pandemic-related events. Interestingly, the percentage of the pro-vaccine group is lower in the Southeast part of the United States. Using multinomial logistic regression, we compare demographics, social capital, income, religious status, political affiliations, geo-locations, sentiment of personal pandemic experience and non-pandemic experience, and county-level pandemic severity perception of these three groups to investigate the scope and causes of public opinions on vaccines. We find that socioeconomically disadvantaged groups are more likely to hold polarized opinions on potential COVID-19 vaccines. The anti-vaccine opinion is the strongest among the people who have the worst personal pandemic experience. Next, by conducting counterfactual analyses, we find that the U.S. public is most concerned about the safety, effectiveness, and political issues regarding potential vaccines for COVID-19, and improving personal pandemic experience increases the vaccine acceptance level. We believe this is the first large-scale social media-based study to analyze public opinions on potential COVID-19 vaccines that can inform more effective vaccine distribution policies and strategies.

---

Researchers suggest that the transmission of SARS-CoV-2 will quickly rebound if interventions (e.g., quarantine and social distancing) are relaxed (Ferguson et al. 2020). Vaccination has greatly reduced the burden of many infectious diseases (Andre et al. 2008) throughout history, and developing SARS-CoV-2 vaccines that can be used globally is, therefore, a priority for ending the pandemic (Yamey et al. 2020). Nevertheless, as scientists and medical experts around the world are developing and testing COVID-19 vaccines, the U.S. public is now divided over whether or not to obtain COVID-19 vaccines. According to a recent Pew Research Center study^1^, in May 71% of U.S. adults indicated that they would definitely or probably obtain a vaccine to prevent COVID-19 if it were available. The percentage dropped sharply, however, to 51% in September. The survey shows that the U.S. public is concerned about the safety and effectiveness of possible vaccines, and the pace of the approval process.

Previous studies show that the sharing of public concerns about vaccines might lead to delaying or not getting vaccination (Gust et al. 2008), which could compromise global COVID-19 vaccine distribution strategies. This phenomenon is termed “vaccine hesitancy” (Dubé et al. 2013) which is a complex issue driven by a variety of context-specific factors (Larson et al. 2014). Researchers have investigated public opinions on existing vaccines for vaccine-preventable diseases like MMR (Motta, Callaghan, and Sylvester 2018; Deiner et al. 2019), HPV (Abdelmutti and Hoffman-Goetz 2010) and H1N1 (Henrich and Holmes 2011). Hesitancy and opinions can vary, however, according to the vaccine involved (Bedford and Lansley 2007). Lazarus et al. (2020) and Feleszko et al. (2020) have investigated the potential acceptance of a COVID-19 vaccine using survey methods, yet little is known about the scope and causes of public opinions on potential COVID-19 vaccines on social media platforms. Meanwhile, the development and testing of COVID-19 vaccines has drawn great attention and response on social media platforms like Twitter and Reddit that allow fast sharing of health information (Scanfeld, Scanfeld, and Larson 2010) and are found to play a major role in disseminating information about vaccinations (Stahl et al. 2016; Dunn et al. 2017). Public attitudes towards the vaccines, therefore, can be reflected by analyzing comments and posts in social media (Kim, Han, and Seo 2020; Tomeny, Vargo, and El-Toukhy 2017).

In the current study, we adopt a human-guided machine learning framework based on state-of-the-art transformer language models to capture individual opinions on potential of COVID-19 vaccines, and categorize these opinions into three groups: pro-vaccine, vaccine-hesitant, anti-vaccine. We use more than 40,000 rigorously selected tweets (out of over six million tweets collected using keywords) posted by over 20,000 distinct Twitter users ranging from September to November of 2020. We aggregate the tweets to reflect the state-level and the national attitudes towards potential COVID-19 vaccines. To characterize the opinion groups, we extract and infer individual-level features such as demographics, social capital, income, religious status, family status, political affiliations, and geo-locations. Lazarus et al. (2020) suggested that personal experience such as COVID-19 sickness in the people and their family, and the external perception such as cases and mortality per million of a nation’s population are associated with the vaccine acceptance level. To quantitatively measure and confirm these two effects, we extract the sentiment of personal pandemic experience and non-pandemic experience for each Twitter user. We collect the number of COVID-19 daily confirmed cases from the data repository maintained by the Center for Systems Science and Engineering (CSSE) at Johns Hopkins University to measure the county-level pandemic severity perception. In our study, we hypothesize that:

- **Hypothesis 1:** There will be differences in demographics, social capital, income, religious status, family status, political affiliations and geo-locations among opinion groups.
- **Hypothesis 2:** The personal pandemic experience will have an impact on shaping the attitude towards potential COVID-19 vaccines.
- **Hypothesis 3:** The county-level pandemic severity perception will have an impact on shaping the attitude towards potential COVID-19 vaccines.

We conduct multinomial logistic regression and find that there are differences in demographics, social capital, income, religious status, political affiliations and geo-locations among the opinion groups. The anti-vaccine opinion is the strongest among the people who have the worst personal pandemic experience. The vaccine-hesitancy is the strongest in the areas that have the worst pandemic severity perceptions. We further show that the individual-level features can be used to anticipate whether this person is in favor of the potential COVID-19 vaccines - or not - over time. By incorporating the individual-level features and additional factor indicators, and by conducting counterfactual analyses, we find that the U.S. public is most concerned about the safety, effectiveness, and political issues with regard to potential vaccines for COVID-19 and improving personal pandemic experience increases the vaccine acceptance level.

## Human-guided machine learning framework

We annotate the opinions of the tweets as pro-vaccine, vaccine-hesitant, or anti-vaccine using a human-guided machine learning framework to strike the best balance between automation and accuracy. In total, we stream over six million publicly available tweets from Twitter using Tweepy API between September 28 to November 4, 2020 with search keywords that are vaccine-related or COVID-19 vaccine-related. Unlike (Tomeny, Vargo, and El-Toukhy 2017), a majority of the tweets crawled with the search keywords in our study is irrelevant to the actual individual opinions about the potential vaccines for COVID-19, which causes a challenging class imbalance problem that may not only slow down the annotation process but also hinder the performance of automated classifiers (Japkowicz and Stephen 2002). To address this problem, we adopt a human-guided machine learning framework (Sadilek et al. 2013) based on the state-of-the-art transformer language model to label the opinions of the tweets. After extracting or inferring the features of these tweets and their authors, we only keep the ones with all the required informative features available.

We initialize the human-guided machine learning framework by sampling 2,000 unique tweets from the corpus *C* with 244,049 tweets. Three researchers independently read each tweet and make a judgement whether this tweet is irrelevant, pro-vaccine, vaccine-hesitant, or anti-vaccine. The label of the tweet is assigned with the consensus votes from three researchers. If three researchers vote entirely differently, the senior researcher determines the label of this tweet after discussing with the other two researchers. The corpus *C*_*train*_ of the initial 2,000 labelled tweets is fed to the XL-Net model (Yang et al. 2019). The four-class classification model *H*_1_ is trained and validated on an external validation set *D*_*validation*_ with 400 annotated tweets^2^. We then construct another binary classification model *H*_2_ that is trained with only two classes of data. One class includes all the irrelevant tweets and the other includes all the relevant tweets that are composed of the pro-vaccine, vaccine-hesitant, and anti-vaccine ones. After training, *H*_2_ is used to make estimates for a corpus of 4,500 unlabelled tweets sampled from *C* regarding whether they are irrelevant or relevant. 90% of a new batch of corpus is composed of the top 10% of the most likely relevant tweets. The other 10% of the new batch is sampled uniformly at random to increase diversity. This new batch of corpus of 500 tweets is annotated by the three researchers as aforementioned and is added to the corpus *C*_*train*_. *H*_1_ is trained with the updated *C*_*train*_ and validated again. This whole process is considered as one iteration.

This framework actively searches for relevant tweets to increase the sizes of the relevant datasets. Figure 1 shows the percentages of the different opinion groups of the original *C*_*train*_ and the final *C*_*train*_ after five iterations. In each iteration, humans guide the machine to learn the irrelevant, pro-vaccine, vaccine-hesitant, and anti-vaccine tweets by updating the training set. Figure 2 shows the performance of *H*_1_ of each iteration. As a result, the framework allows us to label the opinions of the tweets and build the model more efficiently.

**Figure 1:**
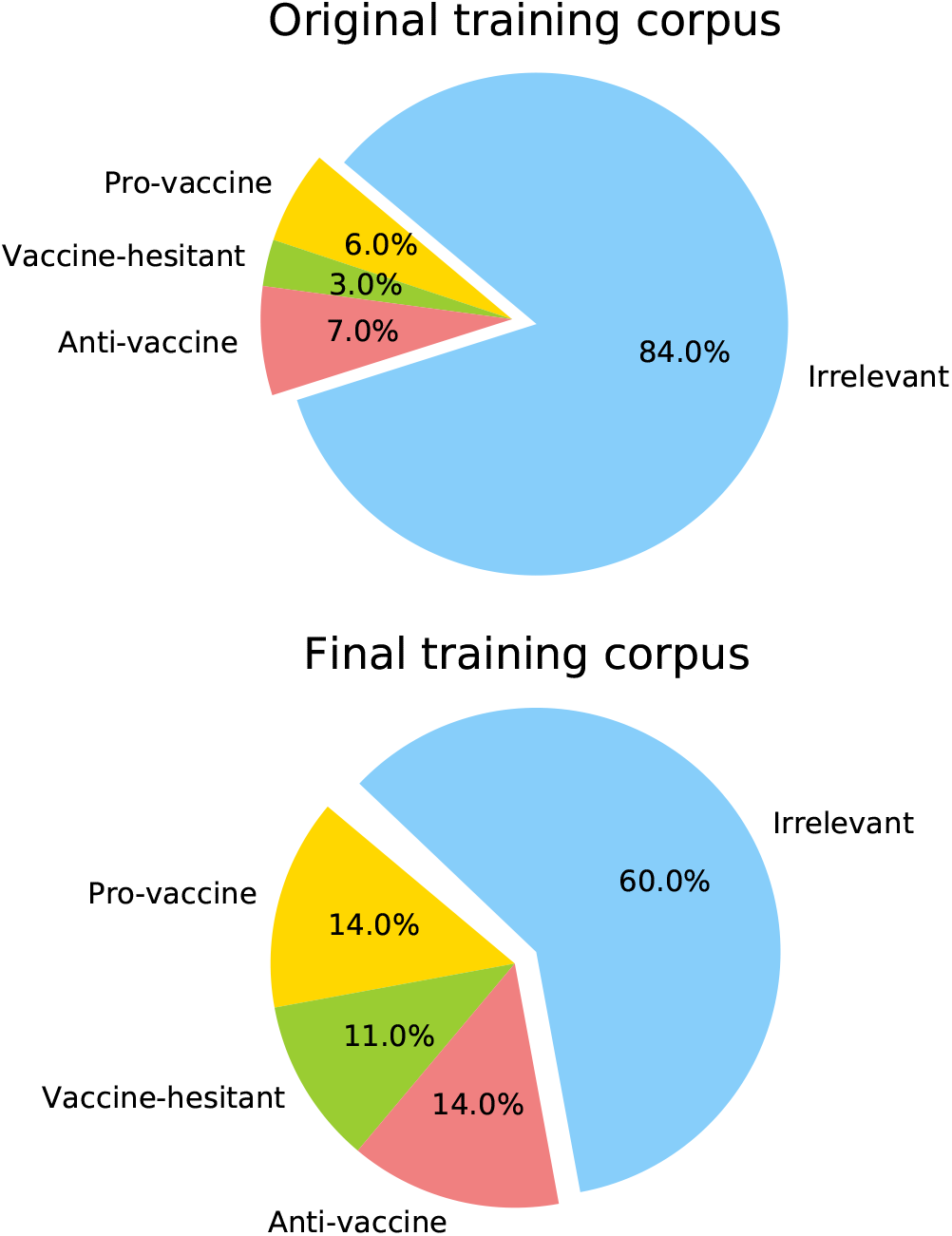
Distributions of different categories of the original and final training corpora.

**Figure 2:**
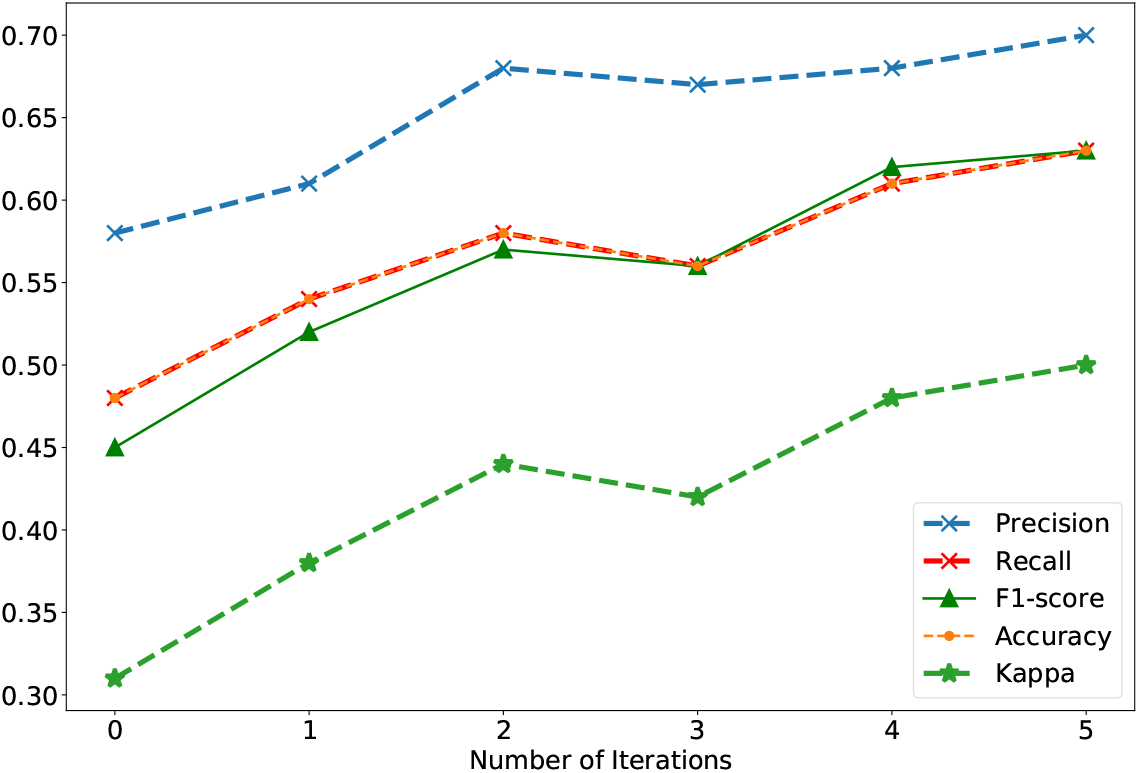
Performance of *H*_1_ of each iteration.

## National and state-level public opinions

The proportions of the different opinion groups of the U.S public change over time as shown in Figure 3, which roughly correspond to the major pandemic-related events. Overall, 57.65% (14,647 of 25,407) are pro-vaccine, 19.30% (4,903 of 25,407) are vaccine-hesitant, and the rest are anti-vaccine. By aggregating people at the state level, we estimate the opinions about the potential COVID-19 vaccines of each state as shown in Figure 4. The Southeast of the U.S. shows a relatively lower acceptance level, so does the cluster of Ohio, Indiana and Kentucky.

**Figure 3:**
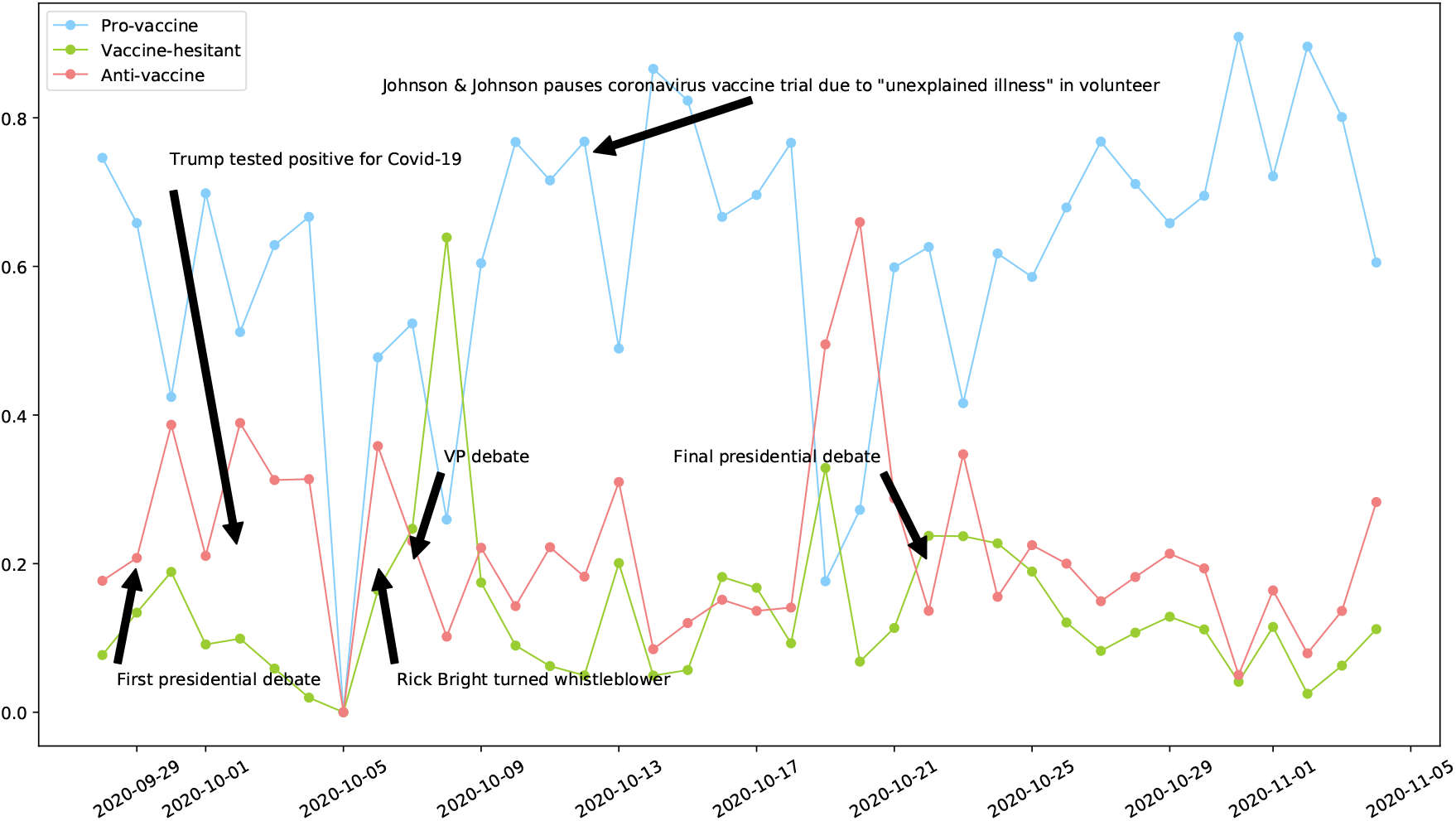
The proportions of the opinion groups from September 28 to November 4, 2020. The data of October 5, 2020 is actually missing due to a data collection issue.

**Figure 4:**
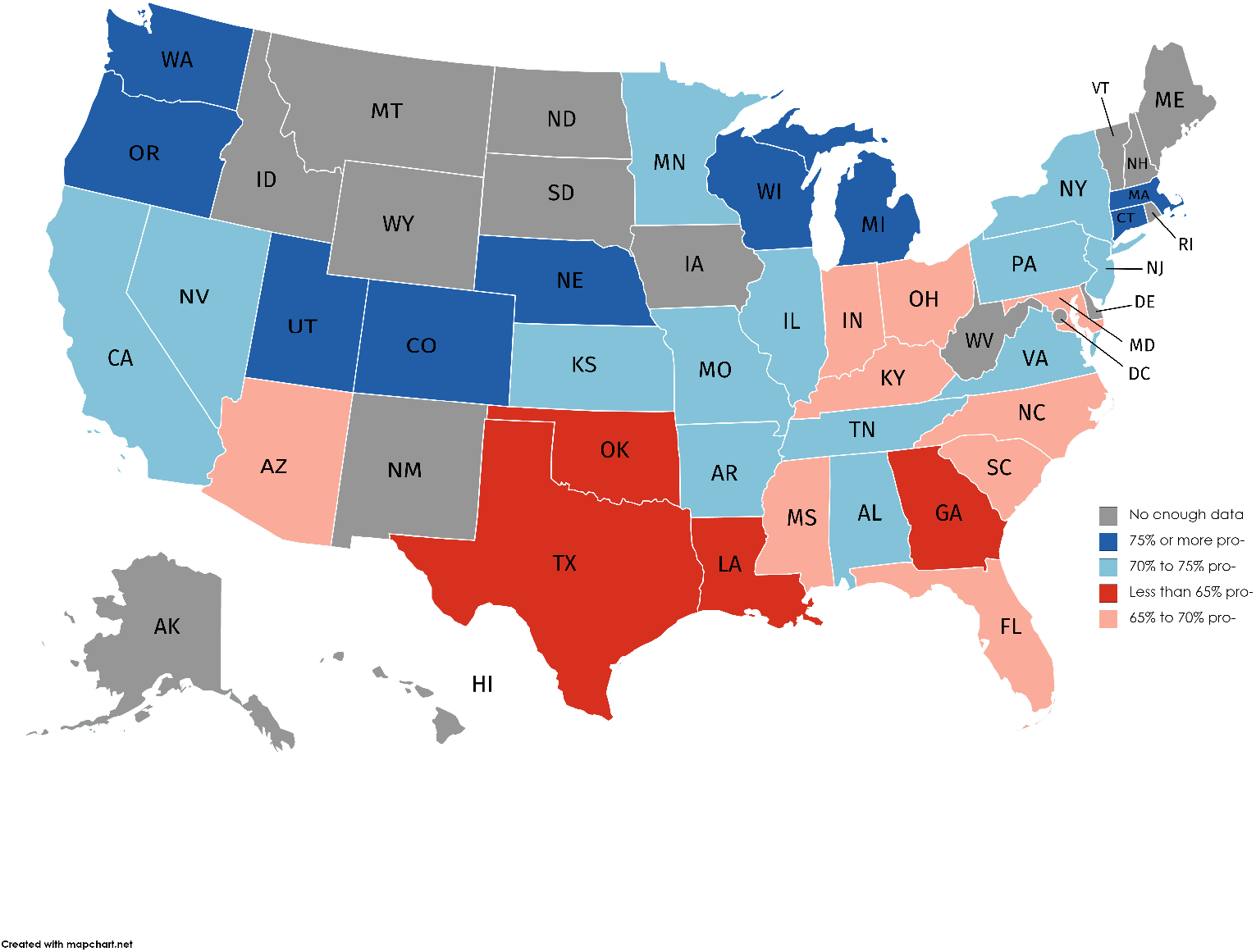
State-level public opinions about potential COVID-19 vaccines.

After performing the Granger Causality Test with a oneday lag, we find that, in Nevada, Tennessee and Washington, the percentage of the pro-vaccine people deviates the most from the national average (*p >* .05). The percentage of the pro-vaccine group of Washington is above the national average during the most of the time, while the acceptance level of Nevada is relatively lower than the national average. More drastic changes are observed for the acceptance level of Tennessee.

## Characterization of different opinion groups

To understand what opinion (i.e., pro-vaccine, vaccine-hesitant, and anti-vaccine) the people (*n* = 10, 945) would hold based on the demographics, social capital, income, religious status, family status, political affiliations, geo-location, sentiment about COVID-19-related experience and non-COVID-related experience, and relative change of the number of daily confirmed cases at the county level, we conduct the multinomial logistic regression, selecting vaccine-hesitant group as the reference category. Descriptive statistics and bi-variate correlations are shown in Table 1. Table 2 summarizes the results of the multinomial logistic regression. The Chi-square test shows that the variables significantly predict the opinion on potential COVID-19 vaccines: *χ*^2^(40, *N* = 10, 945) = 1, 341.49, *p <* .001, Cox and Snell pseudo *R*^2^ = .12, which supports our hypotheses. Next, we show the predictive effects of these variables with paired comparisons.

**Table 1:** Descriptive statistics and the bi-variate correlations. The numbers of `followers`, `friends`, `listed memberships`, `favorites`, `statuses` are normalized by the months of Twitter history and log-transformed.

| Variables | Mean | SD | 1 | 2 | 3 | 4 | 5 | 6 | 7 | 8 | 9 | 10 | 11 | 12 | 13 | 14 | 15 | 16 | 17 | 18 | 19 |
| --- | --- | --- | --- | --- | --- | --- | --- | --- | --- | --- | --- | --- | --- | --- | --- | --- | --- | --- | --- | --- | --- |
| 1. Gender (0 = female, 1 = male) | 0.54 | 0.50 |  |  |  |  |  |  |  |  |  |  |  |  |  |  |  |  |  |  |  |
| 2. Age (years) | 39.89 | 14.69 | .08** |  |  |  |  |  |  |  |  |  |  |  |  |  |  |  |  |  |  |
| 3. Verified (0 = no, 1 = yes) | 0.04 | 0.20 | .00 | .03** |  |  |  |  |  |  |  |  |  |  |  |  |  |  |  |  |  |
| 4. Twitter history (months) | 91.30 | 43.47 | .02 | .03** | .09** |  |  |  |  |  |  |  |  |  |  |  |  |  |  |  |  |
| 5. # Followers | 1.60 | 1.63 | -.04** | -.01 | .37** | -.08** |  |  |  |  |  |  |  |  |  |  |  |  |  |  |  |
| 6. # Friends | 1.95 | 1.25 | -.04** | -.00 | .09** | -.29** | .68** |  |  |  |  |  |  |  |  |  |  |  |  |  |  |
| 7. # Listed memberships | -1.62 | 0.93 | .00 | .08** | .49** | .22** | .69** | .31** |  |  |  |  |  |  |  |  |  |  |  |  |  |
| 8. # Favorites | 4.17 | 1.95 | -.12** | -.14** | -.03** | -.21** | .38** | .47** | .05** |  |  |  |  |  |  |  |  |  |  |  |  |
| 9. # Statuses | 4.09 | 1.43 | -.02* | -.11** | .07** | -.09** | .53** | .45** | .29** | .58** |  |  |  |  |  |  |  |  |  |  |  |
| 10. Higher-Income (0 = no, 1 = yes) | 0.00 | 0.05 | .00 | -.03** | .03** | -.01 | .05** | .02* | .04** | .02** | .01 |  |  |  |  |  |  |  |  |  |  |
| 11. Lower-Income (0 = no, 1 = yes) | 0.76 | 0.43 | .00 | -.43** | -.04** | .05** | -.15** | -.18** | -.12** | -.13** | -.09** | -.10** |  |  |  |  |  |  |  |  |  |
| 12. Religious (0 = no, 1 = yes) | 0.04 | 0.19 | -.01 | .07** | -.03** | -.03** | .03** | .07** | -.03** | .02 | .02 | -.00 | -.06** |  |  |  |  |  |  |  |  |
| 13. Having kids (0 = no, 1 = yes) | 0.12 | 0.32 | -.09** | .09** | .03** | .02* | .04** | .05** | .04** | .01 | -.05** | -.01 | -.05** | .09** |  |  |  |  |  |  |  |
| 14. Following Trump (0 = no, 1 = yes) | 0.11 | 0.31 | .05** | .06** | -.03** | -.17** | -.05** | .04** | -.12** | -.01 | -.05** | -.00 | -.05** | .06** | .04** |  |  |  |  |  |  |
| 15. Following Biden (0 = no, 1 = yes) | 0.17 | 0.38 | -.09** | .07** | .06** | .10** | .05** | .19** | .06** | .09** | .03** | .00 | -.06** | -.03** | .07** | .01 |  |  |  |  |  |
| 16. Rural (0 = no, 1 = yes) | 0.19 | 0.40 | .02 | .09** | -.05** | -.04** | -.07** | -.02 | -.08** | -.01 | -.04** | .00 | -.05** | .06** | .04** | .05** | -.00 |  |  |  |  |
| 17. Suburban (0 = no, 1 = yes) | 0.14 | 0.35 | .01 | .07** | -.01 | -.02* | -.05** | -.03** | -.05** | -.03** | -.02** | -.02 | -.01 | .03** | .04** | .03** | -.00 | -.20** |  |  |  |
| 18. Pandemic experience (sentiment) | 0.06 | 0.80 | -.03** | -.04** | .10** | .03** | .09** | -.00 | .15** | -.08** | -.08** | .02* | .04** | -.03** | -.00 | -.06** | .01 | -.03** | .00 |  |  |
| 19. Non-pandemic experience (sentiment) | 0.62 | 0.75 | -.07** | -.06** | .07** | .04** | .14** | .09** | .14** | .06** | .01 | .02 | .02 | .00 | .03** | -.06** | .01 | -.03** | -.02* | .27** |  |
| 20. Pandemic severity perception (relative change of # daily confirmed cases) | 0.01 | 0.00 | .01 | .03** | -.03** | -.02* | -.05* | -.01 | -.07** | -.00 | -.04** | .01 | -.03** | .06** | .05** | .04** | -.01 | .27** | .12** | -.01 | .01 |

Note. \* $p < 0.05$ . \*\* $p < 0.01$ .
Note. \* $p < 0.05$ . \*\* $p < 0.01$ .

**Table 2:** Multinomial logistic regression outputs for the opinion on potential COVID-19 vaccines against demographics and other variables of interest. The vaccine-hesitant group is selected as the reference category.

| Predictor | Anti-vaccine |  |  | Pro-vaccine |  |  |
| --- | --- | --- | --- | --- | --- | --- |
|  | *B* | *SE* | OR (95% CI) | *B* | *SE* | OR (95% CI) |
| Intercept | -1.80*** | 0.26 |  | 0.79*** | 0.20 |  |
| Age (years) | 0.00 | 0.00 | 1.00 (0.995, 1.005) | 0.01*** | 0.00 | 1.01 (1.01, 1.02) |
| Twitter history (months) | -0.001 | 0.001 | 0.999 (0.997, 1.000) | -0.003*** | 0.001 | 0.997 (0.996, 0.999) |
| # Followers | 0.28*** | 0.04 | 1.32 (1.22, 1.42) | 0.08** | 0.03 | 1.09 (1.02, 1.16) |
| # Friends | -0.18*** | 0.04 | 0.83 (0.77, 0.90) | -0.07* | 0.03 | 0.93 (0.88, 0.99) |
| # Listed memberships | -0.62*** | 0.06 | 0.54 (0.48, 0.61) | 0.10* | 0.04 | 1.11 (1.01, 1.20) |
| # Favorites | 0.04* | 0.02 | 1.05 (1.00, 1.09) | 0.04* | 0.02 | 1.04 (1.01, 1.08) |
| # Statuses | 0.11*** | 0.03 | 1.12 (1.06, 1.19) | -0.06* | 0.02 | 0.94 (0.90, 0.99) |
| Pandemic experience (sentiment) | -0.18*** | 0.04 | 0.84 (0.77, 0.91) | 0.21*** | 0.03 | 1.24 (1.16, 1.32) |
| Non-pandemic experience (sentiment) | -0.04 | 0.04 | 0.97 (0.89, 1.04) | 0.13*** | 0.03 | 1.14 (1.07, 1.22) |
| Pandemic severity perception<br>(relative change of # daily confirmed cases) | -14.91 | 8.13 | 0.00 (0.00, 2.78) | -22.68*** | 6.59 | 0.00 (0.00, 0.00) |
| Female | -0.25*** | 0.06 | 0.78 (0.69, 0.88) | -0.47*** | 0.05 | 0.63 (0.57, 0.69) |
| Verified user | -0.63* | 0.27 | 0.53 (0.32, 0.90) | -0.16 | 0.14 | 0.85 (0.65, 1.12) |
| Higher-income | -20.76*** | 0.00 | 0.00 (0.00, 0.00) | 0.47 | 0.43 | 1.60 (0.70, 3.67) |
| Lower-income | 0.40*** | 0.08 | 1.49 (1.26, 1.75) | 0.52*** | 0.06 | 1.69 (1.49, 1.91) |
| Religious | 0.74*** | 0.17 | 2.10 (1.52, 2.91) | 0.37* | 0.15 | 1.45 (1.08, 1.95) |
| Having kids | -0.11 | 0.10 | 0.90 (0.74, 1.09) | -0.00 | 0.08 | 1.00 (0.86, 1.16) |
| Following Trump | 0.42*** | 0.10 | 1.52 (1.26, 1.83) | 0.06 | 0.08 | 1.06 (0.90, 1.25) |
| Following Biden | -1.22*** | 0.10 | 0.30 (0.25, 0.36) | -0.34*** | 0.06 | 0.71 (0.63, 0.80) |
| Rural | 0.17* | 0.08 | 1.19 (1.01, 1.39) | 0.07 | 0.07 | 1.07 (0.94, 1.22) |
| Suburban | 0.19* | 0.09 | 1.20 (1.01, 1.44) | 0.11 | 0.07 | 1.12 (0.97, 1.29) |
| Chi-square | 1,341.49*** |  |  |  |  |  |
| *df* | 40 |  |  |  |  |  |
| -2 log likelihood | 20,171.34 |  |  |  |  |  |
| Cox and Shell pseudo $R^2$ | 0.12 | | | | | |
| Sample size | 10,945 |  |  |  |  |  |

Note. \* $p < 0.05$ . \*\* $p < 0.01$ . \*\*\* $p < 0.001$ .

### Female are more likely to hold hesitant opinions

Gender is statistically significant (*χ*^2^ = 91.83, *p <* .001). Females are likely to hold hesitant opinions rather than polarized opinions (i.e., pro-vaccine, anti-vaccine). Specifically, comparing the anti-vaccine group and vaccine-hesitant group, we find that females are less likely to be anti-vaccine (*B* = *−*0.25, *SE* = 0.06, *p <* .001, *OR* = 0.78; 95%*CI* = [0.69, 0.88]). Comparing the pro-vaccine group and vaccine-hesitant group, we find that females are also less likely to be pro-vaccine (*B* = *−*0.47, *SE* = 0.05, *p <* .001, *OR* = 0.63; 95%*CI* = [0.57, 0.69]).

### Older people tend to be pro-vaccine

Age is statistically significant (*χ*^2^ = 72.47, *p <* .001). Comparing the anti-vaccine group and vaccine-hesitant group, we do not find significant evidence that older people are more anti-vaccine. However, comparing the pro-vaccine group and vaccine-hesitant group, we find that people who are one year older are 1.01 (*B* = 0.01, *SE* = 0.00, *p <* .001, *OR* = 1.01; 95%*CI* = [1.01, 1.02]) times more likely to be pro-vaccine instead of vaccine-hesitant, which echoes the study of Lazarus et al. (2020). One potential explanation is that the risk of dying with COVID-19 increases with age (Lloyd-Sherlock et al. 2020), and the benefits of not getting infected with COVID-19 outweigh the risk of getting vaccinated.

### Different patterns of Twitter usage

A `Verified` Twitter account must represent or other wise be associated with a prominently recognized individual or brand^3^. In our study, `Verified` status is statistically significant (*χ*^2^ = 6.12, *p <* .05). Comparing the anti-vaccine group and vaccine-hesitant group, we find `Verified` users are less likely to be anti-vaccine (*B* = *−*0.63, *SE* = 0.27, *p <* .05, *OR* = 0.53; 95%*CI* = [0.32, 0.90]), however, comparing the pro-vaccine group and vaccine-hesitant group, we do not find significant differences.

Months of Twitter history is statistically significant (*χ*^2^ = 17.52, *p <* .001). Comparing the anti-vaccine group and vaccine-hesitant group, we do not find significant differences, however, comparing the pro-vaccine group and vaccine-hesitant group, we find if the months of Twitter history were to increase by one month, the probability for being pro-vaccine rather than vaccine-hesitant would be expected to be decreased by a factor 0.997 (*B* = *−*0.003, *SE* = 0.001, *p <* .001, *OR* = 0.997; 95%*CI* = [0.996, 0.999]).

After normalizing the number of `followers`, `friends`, `listed memberships`, `favorites`, and `statuses` with the number of months of Twitter history, we still find that the social capital is statistically significant. Specifically, there are significant differences in terms of `followers` counts (*χ*^2^ = 51.06, *p <* .001), `friends` counts (*χ*^2^ = 21.28, *p <* .001), `listed memberships` counts (*χ*^2^ = 199.51, *p <* .001), `favorites` counts (*χ*^2^ = 6.10, *p <* .05), `statuses` counts (*χ*^2^ = 47.37, *p <* .001).

Comparing the anti-vaccine group and vaccine-hesitant group, if the log-scale `followers` count were to increase by one unit, it is 1.32 (*B* = 0.28, *SE* = 0.04, *p <* .001, *OR* = 1.32; 95%*CI* = [1.22, 1.42]) times more likely to be anti-vaccine. If the log-scale `friends` count were to increase by one unit, it is less likely to be anti-vaccine (*B* = *−*0.18, *SE* = 0.04, *p <* .001, *OR* = 0.83; 95%*CI* = [0.77, 0.90]). If the log-scale `listed memberships` count were to increase by one unit, it is less likely to be anti-vaccine (*B* = *−*0.62, *SE* = 0.06, *p <* .001, *OR* = 0.54; 95%*CI* = [0.48, 0.61]). If the log-scale `favorites` count were to increase by one unit, it is 1.05 (*B* = 0.04, *SE* = 0.02, *p <* .05, *OR* = 1.05; 95%*CI* = [1.00, 1.09]) times more likely to be anti-vaccine. If the log-scale `statuses` count were to increase by one unit, it is 1.12 (*B* = 0.11, *SE* = 0.03, *p <* .001, *OR* = 1.12; 95%*CI* = [1.06, 1.19]) times more likely to be anti-vaccine.

Comparing the pro-vaccine group and vaccine-hesitant group, if the log-scale `followers` count were to increase by one unit, it is 1.09 (*B* = 0.08, *SE* = 0.03, *p <* .01, *OR* = 1.09; 95%*CI* = [1.02, 1.16]) times more likely to be pro-vaccine. If the log-scale `friends` count were to increase by one unit, it is less likely to be pro-vaccine (*B* = *−*0.07, *SE* = 0.03, *p <* .05, *OR* = 0.93; 95%*CI* = [0.88, 0.99]). If the log-scale `listed memberships` count were to increase by one unit, it is 1.11 (*B* = 0.10, *SE* = 0.04, *p <* .05, *OR* = 1.11; 95%*CI* = [1.01, 1.20]) times more likely to be pro-vaccine. If the log-scale `favorites` count were to increase by one unit, it is 1.04 (*B* = 0.04, *SE* = 0.02, *p <* .05, *OR* = 1.04; 95%*CI* = [1.01, 1.08]) times more likely to be pro-vaccine. If the log-scale `statuses` count were to increase by one unit, it is less likely to be pro-vaccine (*B* = *−*0.06, *SE* = 0.02, *p <* .05, *OR* = 0.94; 95%*CI* = [0.90, 0.99]).

Twitter users who have more `followers` or fewer `friends`, or give more `favourites` are more likely to hold polarized opinion. The larger `listed memberships` count is, the more likely the Twitter user is pro-vaccine. Twitter users who post more `statuses` tend to be anti-vaccine.

### The lower-income group is more likely to hold polarized opinions

Income is statistically significant (*χ*^2^ = 79.09, *p <* .001). Comparing the anti-vaccine group and vaccine-hesitant group, we find that the lower-income group is 1.49 (*B* = 0.40, *SE* = 0.08, *p <* .001, *OR* = 1.49; 95%*CI* = [1.26, 1.75]) times more likely to be anti-vaccine than the medium-income group, and the higher-income group is less likely to be anti-vaccine than the medium-income group (*B* = *−*20.76, *SE* = 0.00, *p <* .001, *OR* = 0.00; 95%*CI* = [0.00, 0.00]). Comparing the pro-vaccine group and vaccine-hesitant group, we find that lower-income group is 1.69 (*B* = 0.52, *SE* = 0.06, *p <* .001, *OR* = 1.69; 95%*CI* = [1.49, 1.91]) times more likely to be pro-vaccine than medium-income group. The difference between the higher-income group and medium-income group is not significant. *Inconsistent* with Lazarus et al. (2020) that the higher the income is, the more likely people are pro-vaccine, we find the effect of income more nuanced. Lower-income people tend to be polarized.

### Religious people are more likely to be polarized

Religious status is statistically significant (*χ*^2^ = 21.34, *p <* .001). Comparing the anti-vaccine group and vaccine-hesitant group, we find that religious people are more likely to be anti-vaccine than non-religious people (*B* = 0.74, *SE* = 0.17, *p <* .001, *OR* = 2.10; 95%*CI* = [1.52, 2.91]). Comparing the pro-vaccine group and vaccine-hesitant group, we find that religious people are also more likely to be pro-vaccine than religious people (*B* = 0.37, *SE* = 0.15, *p <* .05, *OR* = 1.45; 95%*CI* = [1.08, 1.95]). This is in line with Larson et al. (2014) that the effect of religious status is complicated.

### Political diversion indicates a divided opinion about the potential COVID-19 vaccines

Following Donald Trump is statistically significant (*χ*^2^ = 25.22, *p <* .001). Comparing the anti-vaccine group and vaccine-hesitant group, we find that the Twitter users who follow Donald Trump are 1.52 (*B* = 0.42, *SE* = 0.10, *p <* .001, *OR* = 1.52; 95%*CI* = [1.26, 1.83]) times more like to be anti-vaccine than the Twitter users who do not. Comparing the pro-vaccine group and vaccine-hesitant group, following Donald Trump is not significant.

Following Joe Biden is statistically significant (*χ*^2^ = 177.96, *p <* .001). Comparing the anti-vaccine group and vaccine-hesitant group, we find that the Twitter users who follow Joe Biden are less like to be anti-vaccine than the Twitter users who do not (*B* = *−*1.22, *SE* = 0.10, *p <* .001, *OR* = 0.30; 95%*CI* = [0.25, 0.36]). Comparing the pro-vaccine group and vaccine-hesitant group, we find that the Twitter users who follow Joe Biden are also less likely to be pro-vaccine than the Twitter users who do not (*B* = *−*0.34, *SE* = 0.06, *p <* .001, *OR* = 0.71; 95%*CI* = [0.63, 0.80]).

Twitter users who follow Donald Trump tend to be anti-vaccine, while those who follow Joe Biden tend to be vaccine-hesitant.

### People living in suburban or rural areas are more likely to be anti-vaccine

Although the population density of the area is not statistically significant across three opinion categories, we still find differences between the anti-vaccine group and vaccine-hesitant group. People living in suburban areas are 1.20 (*B* = 0.19, *SE* = 0.09, *p <* .05, *OR* = 1.20; 95%*CI* = [1.01, 1.44]) times more likely to be anti-vaccine than people living in urban areas. People living in rural areas are 1.19 (*B* = 0.17, *SE* = 0.08, *p <* .05, *OR* = 1.18; 95%*CI* = [1.01, 1.39]) times more likely to be anti-vaccine than people living in urban areas.

Most of the results are consistent with Hypothesis 1. There are significant differences in demographics, social capital, income, religious status, political affiliations and geo-locations among opinion groups, however, we do not find significant difference in family status.

### Personal experience with COVID-19 and the county-level pandemic severity perception shape the opinion

The sentiment score of personal experience with COVID-19 is statistically significant (*χ*^2^ = 146.50, *p <* .001). Comparing the anti-vaccine group and vaccine-hesitant group, we find that if the sentiment score of personal experience with COVID-19 were to increase by one unit (i.e., the sentiment became more positive), the person would be less likely to hold anti-vaccine opinion (*B* = *−*0.18, *SE* = 0.04, *p <* .001, *OR* = 0.84; 95%*CI* = [0.77, 0.91]). Comparing the pro-vaccine group and vaccine-hesitant group, we find if the sentiment score of personal experience with COVID-19 were to increase by one unit (i.e., the sentiment became more positive), the person would be 1.24 times more likely to hold pro-vaccine opinion (*B* = 0.21, *SE* = 0.03, *p <* .001, *OR* = 1.24; 95%*CI* = [1.16, 1.32]), which is consistent with Hypothesis 2.

The sentiment score of non-COVID-related personal experience is overall statistically significant (*χ*^2^ = 29.28, *p <* .001), but comparing the anti-vaccine group and vaccine-hesitant group, we find no significant difference. However, comparing the pro-vaccine group and vaccine-hesitant group, we find if the sentiment score of non-COVID-related personal experience were to increase by one unit (i.e., the sentiment became more positive), the person would be more likely to hold pro-vaccine opinion (*B* = 0.13, *SE* = 0.03, *p <* .001, *OR* = 1.14; 95%*CI* = [1.07, 1.22]).

The county-level pandemic severity perceptions are overall statistically significant (*χ*^2^ = 11.76, *p <* .01), supporting Hypothesis 3, but we find no significant difference comparing the anti-vaccine group and vaccine-hesitant group. However, comparing the pro-vaccine group and vaccine-hesitant group, if the relative change of the number of daily confirmed cases at the county level were to increased by one unit, the person would be less likely to be pro-vaccine (*B* = *−*22.71, *SE* = 6.59, *p <* .001, *OR* = 0.00; 95%*CI* = [0.00, 0.00]).

At the individual level, the personal pandemic experience is a strong predictor of the opinion about COVID-19 vaccines. The anti-vaccine opinion is the strongest among the people who have the worst pandemic experience. However, the non-pandemic experience is not a strong predictor of anti-vaccine opinion. At the county level, the vaccine-hesitance is the strongest in the areas that have the worst pandemic severity perception (i.e., the relative change of the number of daily confirmed cases is the largest).

We conduct multinomial logistic regression to investigate the scope and causes of public opinions on vaccines and test three hypotheses. The current study shows the hypothesized effects of most of the characteristics in predicting the odds of being pro-vaccine or anti-vaccine against vaccine-hesitant. The findings suggest that females are more vaccine-hesitant, which is consistent with the Reuters/Ipsos survey^4^, and older people tend to be pro-vaccine. With respect to social capital, people who have more `followers` or fewer `friends`, or give more `favorites`, are more likely to hold polarized opinions. `Verified` status, months of Twitter history, `listed memberships` counts and `statuses` counts are statistically significant as well. We also show that the lower-income group is more likely to hold polarized opinions. This is *inconsistent* with the finding by Lazarus et al. (2020). Moreover, religious people tend to hold polarized opinions. As for political affiliations, Titter users who follow Donald Trump are more likely to be anti-vaccine rather than vaccine-hesitant, while those who follow Joe Biden tend to be vaccine-hesitant rather than anti-vaccine or pro-vaccine. In addition, we find people who live in rural or suburban areas tend to be anti-vaccine. However, we do not find the hypothesized predictive effect of family status on the opinion about vaccines.

Furthermore, the current study shows the hypothesized predictive effects of the personal pandemic experience and the county-level pandemic severity perception. In particular, personal experience with COVID-19 is a strong predictor of anti-vaccine opinion. The more negative the experience is, the more negative the opinion on vaccines is. People are more likely to be vaccine-hesitant if their pandemic severity perceptions are worse.

## Counterfactual analyses

We next use the aforementioned variables to predict the opinion groups of the user whether this user is pro-vaccine, vaccine-hesitant or anti-vaccine. The data ranging from September 28 to October 21, 2020 are used to train a support vector machine (SVM) *H*_3_ which makes predictions about the opinion group of the data of the latest two weeks (October 22 - November 4, 2020). The real percentage of pro-vaccine users and the prediction one are plotted in Figure 7. The real percentage falls within in one standard deviation of the prediction one.

**Figure 5:**
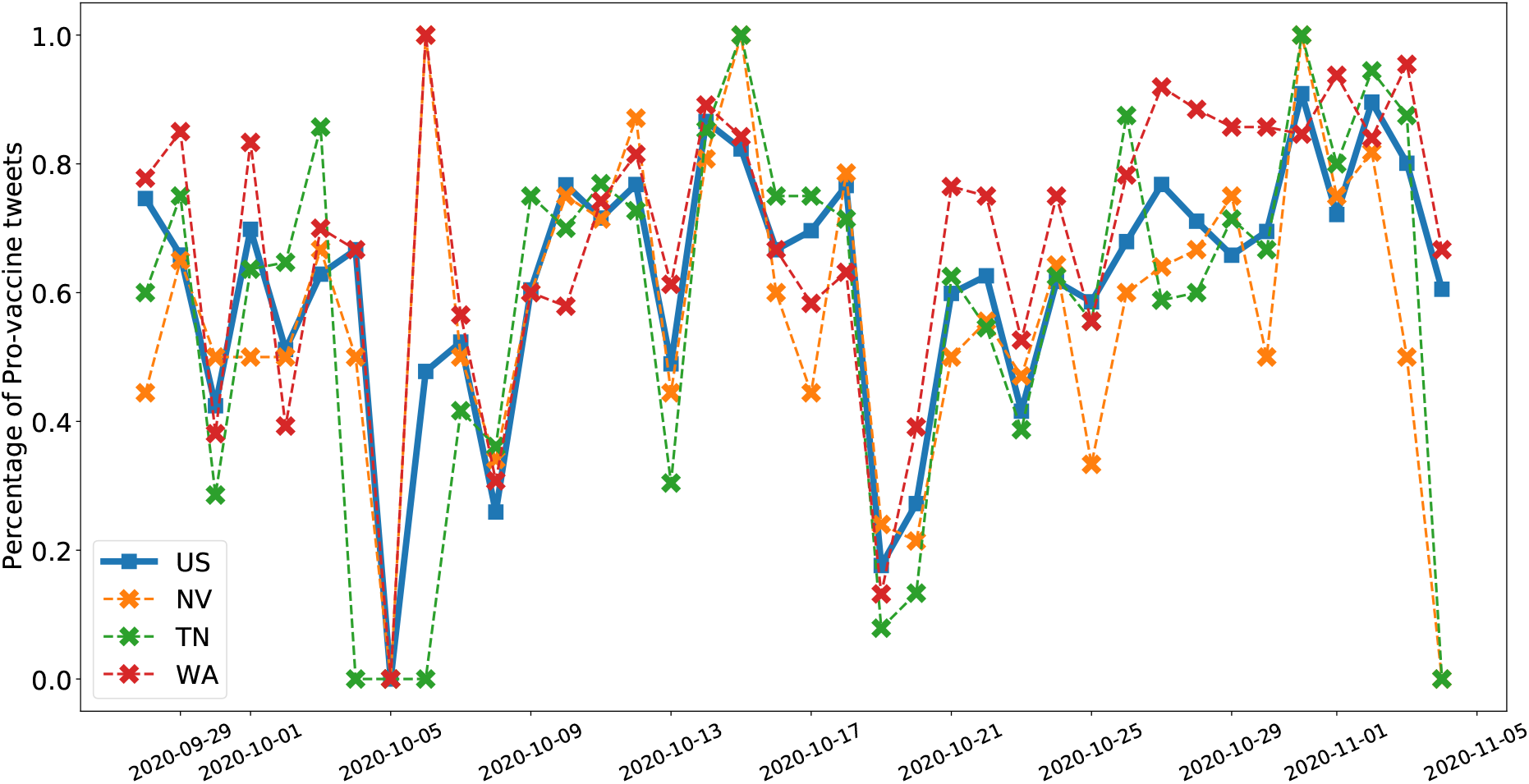
The percentages of the pro-vaccine groups of the national average, Nevada, Tennessee, and Washington.

**Figure 6:**
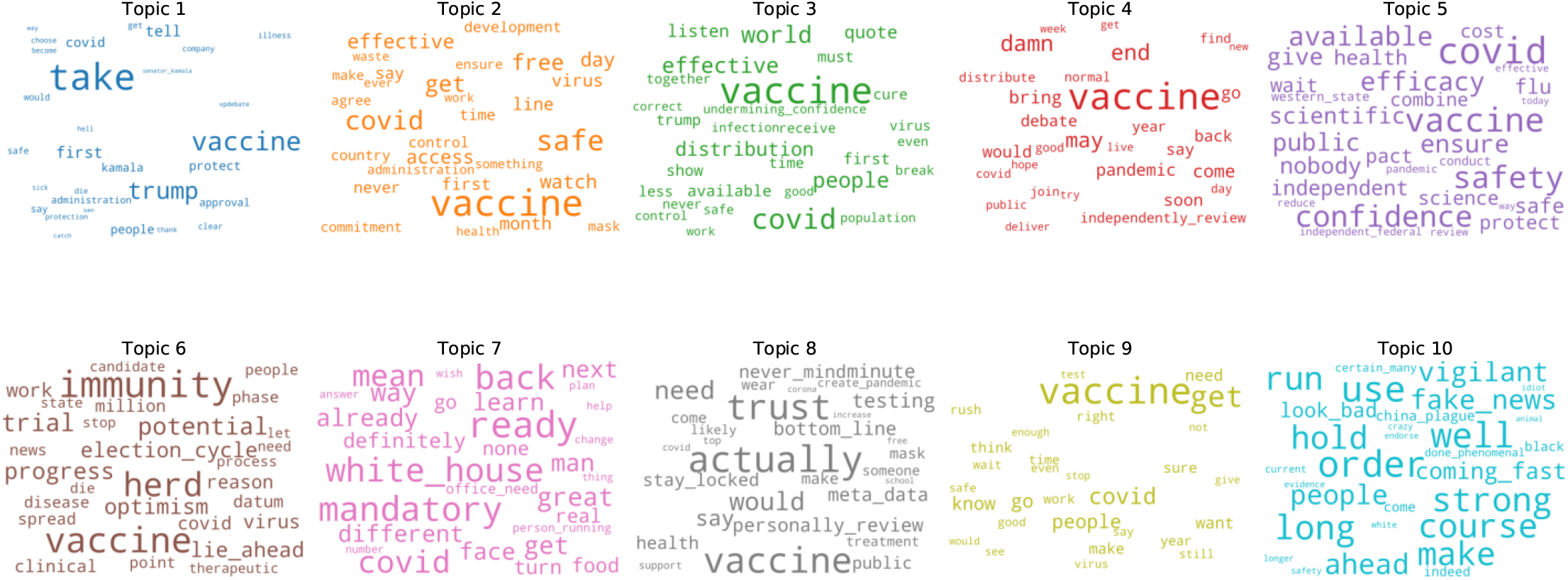
10 topics extracted from the tweets with the top 30 keywords.

**Figure 7:**
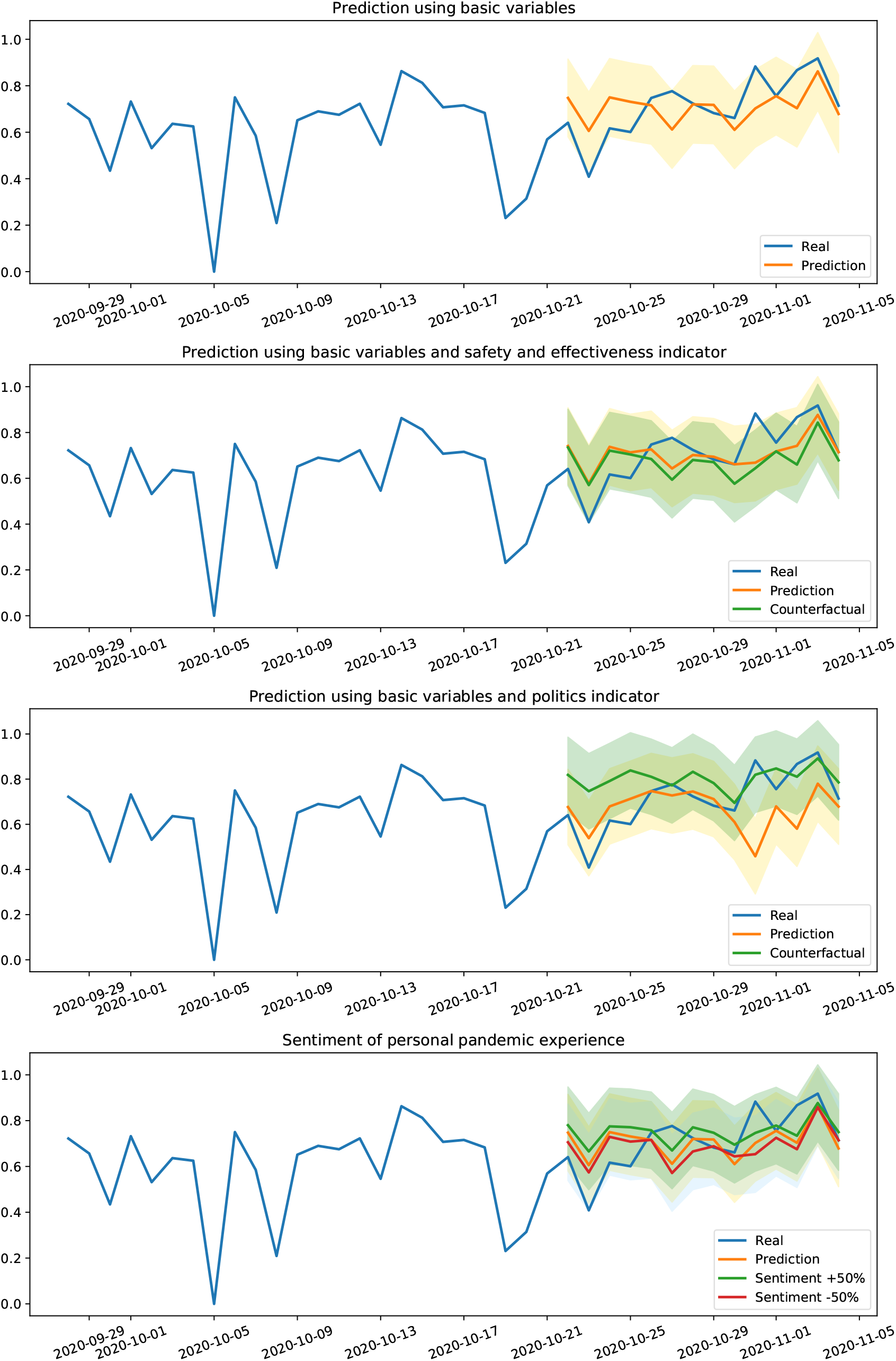
Counterfactual analyses illustrate the importance of politics, safe and effectiveness factor indicators, and personal pandemic experience.

**Figure 8:**
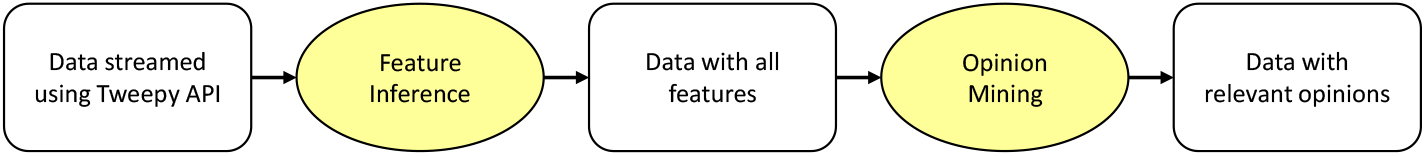
A diagram of data preprocessing procedures.

### Factor indicator

We further analyze the relationship between the opinions and the topics of the tweets using the Latent dirichlet allocation (LDA) topic modelling (Blei, Ng, and Jordan 2003) with 10 topics as shown in Figure 6. In the word cloud of each topic, top 30 keywords are plotted. As we can see from the figure, people are most concerned about the safety and effectiveness of the vaccine which is consistent with the Pew Research Center survey^5^. Some politics-related keywords like “administration”, “white house”, and the names of political figures like “Trump” and “Kamala” are presented as well.

Using counterfactual analyses, we show that our model based on the aforementioned variables and additional factor indicators can inform the distribution policy for the potential COVID-19 vaccines. According to the topics and the keywords extracted by LDA, we narrow down the 10 topics to two major ones: “safety and effectiveness” and “politics”. Using keyword search methods^6^, each tweet is labelled 1 if it contains the related keywords, and 0 if it does not. As a result, except for the basic variables, two more factor indicators are attached. Table 3 shows the descriptions of these two variables. The basic settings for the counterfactual classifiers are the same as *H*_3_. We analyze one factor at a time. We train the classifier with the basic variables and the factor indicator of the original value. The prediction is plotted in orange in Figure 7. Then we change the value of the factor indicator which was originally 1 into 0, keeping other variables constant. The trained classifier is used on the counterfactual data, and the prediction is plotted in green.

**Table 3:** Descriptions of the factor indicators.

| Variables | Yes (%) | No (%) |
| --- | --- | --- |
| 1. Politics | 2,749 (25.12%) | 8,196 (74.88%) |
| 2. Safety and effectiveness | 1,971 (18.01%) | 8,974 (81.99%) |

**Table 4:** Labeling Standards for Tweets

| Category | Description |
| --- | --- |
| Pro-vaccine | i. Claiming that they would take the vaccine once it is available<br>ii. Advocating and supporting vaccine/vaccine-associated entities like vaccine experiment trials<br>iii. Believing that the vaccine will be the solution to the pandemic |
| Vaccine-hesitant | i. Claiming that they would like to take the vaccine given that the vaccine is proven safe/effective<br>ii. Claiming that they would wait for a while and see whether a vaccine is truly safe/effective if there is one<br>iii. Showing worries about the effectiveness of a rushed vaccine regardless of tones |
| Anti-vaccine | i. Promoting/arguing in favor of conspiracy theory about vaccine/vaccine-associated entities<br>ii. Believing that an effective vaccine would not be invented quickly and help us overcome the pandemic<br>iii. Believing that a covid-19 vaccine is dangerous for whatever reasons and would not take it even though the commenters claim that they are not anti-vaccine |
| Irrelevant | i. Vaccine News. No written opinion from the commenters<br>ii. Including vaccine and the commenters’ opinions, but the focus is something else (i.e., insurance, politics, personal life experience, economics, emotional complaints, etc.)<br>iii. Comments/questions on vaccines/vaccine-associated entities but with unclear meanings |

**Table 5:** Performance of the four-class XLNet model *H*_1_

| Class | Precision | Recall | F1-score |
| --- | --- | --- | --- |
| Irrelevant | 0.45 | 0.84 | 0.59 |
| Pro-vaccine | 0.78 | 0.52 | 0.62 |
| Vaccine-hesitant | 0.77 | 0.54 | 0.64 |
| Anti-vaccine | 0.79 | 0.61 | 0.69 |
| Overall | 0.7 | 0.63 | 0.63 |

### Removing the safety and effectiveness factors reduces the vaccine acceptance level. However, removing the politics factor increases it

Figure 7 shows the results of counterfactual analyses of factor indicators. Using counter-factual analysis by turning the factor indicator of safety and effectiveness into 0, there is a clear decrease (4.42% on average) of the percentage of the pro-vaccine people. However, by turning the factor indicator of politics into 0, there is a clear increase (22.65% on average) of the percentage of the pro-vaccine people. This indicates that people are most concerned about the relationship between the politics and the potential COVID-19 vaccines, which is also mirrored by the news report^7^.

### Improving personal pandemic experience increases the vaccine acceptance level

Figure 7 shows the results pf counterfactual analyses of different sentiment levels of personal pandemic experience. By increasing the sentiment scores with a factor of 50%, the percentage of the pro-vaccine people increases by 6.39%. However, by reducing the sentiment scores of a factor of 50%, the percentage of the pro-vaccine people decreases by 2.82%.

## Discussion

Our current study has limitations. The public opinions of some (less populated) states cannot be reflected due to the inadequate data. The findings could be further validated in other populations. However, our study broadly captures the public opinions on the potential vaccines for COVID-19 on Twitter. By aggregating the opinions, we find a lower acceptance level in the Southeast part of the U.S. The changes of the proportions of different opinion groups correspond roughly to the major pandemic-related events. We show the hypothesized predictive effects of the characteristics of the people in predicting pro-vaccine, vaccine-hesitant, and anti-vaccine group. For example, the socioeconomically disadvantaged groups have a relatively more polarized attitude towards the potential vaccines. The personal pandemic experience and the county-level pandemic severity perception shape the opinions. Specifically, the anti-vaccine opinion is the strongest among the people who have the worst personal pandemic experience, and the vaccine-hesitancy is the strongest in the areas that have the worst pandemic severity perception. Using counterfactual analyses, we find that people are most concerned about the safety, effectiveness and politics regarding potential COVID-19 vaccines, and improving personal experience with COVID-19 increases the vaccine acceptance level.

Our results can guide and support policymakers making more effective distribution policies and strategies. First, more efforts of dissemination should be spent on the socioeconomically disadvantaged groups who are exposed to potentially higher risks (Chang et al. 2020; Hopman, Allegranzi, and Mehtar 2020; Adams-Prassl et al. 2020) and already possess more polarized attitudes towards the vaccines. Second, messaging for the vaccines is extremely important because the vaccine acceptance level can be increased by removing the politics factor. Third, safety and effectiveness issues need to be well addressed because the acceptance level is reduced by removing this factor. Finally, improving personal pandemic experience may increase the vaccine acceptance level as well and thus all helpful measures should be integrated to maximize the vaccine acceptance. In the future, by combining social media data and more traditional survey data, we hope to acquire deeper insights into the public opinions on potential COVID-19 vaccines and thus inform more effective vaccine dissemination policies and strategies.

## Data Availability

Data sharing is not applicable.

## Methods

The Methods section is structured as follows. We describe the datasets we use in Methods M1 and how we infer or extract features in Methods M2. We describe our strategy for opinion mining and the standard of labelling in Methods M3. In Methods M4, we discuss the experimental procedures.

### M1 DataSets

#### Twitter

We use the Tweepy API^8^ to collect the related tweets which are publicly available. The search keywords and hashtags are COVID-19 vaccine-related or vaccine-related, including “vaccine”, “COVID-19 vaccine”, “COVID vaccine”, “COVID19 vaccine”, “vaccinated”, “immunization”, “covidvaccine”, “covid19vaccine” and “#vaccine”.^9^ Slang and misspellings of the related keywords are also included which are composed of “vacinne”, “vacine”, “antivax” and “anti vax”. In the end, 6,314,327 tweets (including retweets) from September 28 to November 4, 2020 posted by 1,874,468 distinct Twitter users are collected.

The tweet content and other Twitter profile information are used to extract or predict demographics, user-level features like the number of `followers`, income, religious status, family status, political affiliations, geo-locations, sentiment about the COVID-19-related experience and non-COVID-related experience. To infer the family status, religious status and sentiment, we use Tweepy API to collect the publicly available tweets posted by each user for the last three months. For example, if the tweet containing the search keywords or hashtags was posted on October 1, 2020, then all the publicly available tweets posted by this Twitter user from July 1 to October 1, 2020 are collected as well.

The preprocessing pipeline is shown in Figure 8. First, the features of the Twitter users are inferred or extracted. To better understand the relationships between all characteristics, we choose to only keep the users of which we can infer all the features except for sentiment. Next, we achieve the mining of opinions via a human-guided machine learning framework. In the end, we use 40,210 rigorously selected tweets and 25,407 unique Twitter users to study the national and state-level public opinions. 10,945 unique Twitter users with sentiment of personal pandemic experience and non-pandemic experience are included in the characterization study and counterfactual analyses.

#### JHU CSSE

We extract the number of COVID-19 daily confirmed cases from the data repository maintained by the Center for Systems Science and Engineering (CSSE) at Johns Hopkins University^10^. The median relative change of the number of daily confirmed cases of the last three months at the county level is calculated to measure the county-level pandemic severity perception.

### M2 Feature Inference

#### Demographics

Following the methods of Lyu et al. (2020), we use Face++ API^11^ to infer the gender and age information of the users using their profile images. The invalid image urls and images with multiple or zero faces are excluded. The gender and age information of the remaining users (i.e., there is only one intelligible face in the profile image) is inferred. Since our study focuses on the opinions of U.S. adults, the users who are younger than 18 are removed.

#### User-level features

Seven user-level features are crawled by Tweepy API as well which include the number of `followers`, `friends`, `listed memberships`, `favourites`, `statuses`, the number of months since the user account was created, and the `Verified` status. Moreover, we normalize the number of `followers`, `friends`, `listed memberships`, `favourites`, and `statuses` by the number of months since the user account was created.

#### Geo-locations

For Twitter, we choose to resolve the geo-locations using users’ profiles. Similar to Lyu et al. (2020), the locations with noise are excluded, and the rest are classified into urban, suburban, or rural.

#### Income

We design a supervised ensemble model to predict the income of Twitter users. The ensemble model includes Gradient Boost Decision Tree (GBDT), Random Forest, Logistic Regression, and XGboost. We use the income datasets of Twitter users (Preoţiuc-Pietro et al. 2015) to train our model(s). The features include age, days of Twitter history, the number of `followers`, `friends`, `listed memberships`, `favourites`, and sentiment score calculated by Vader (Gilbert and Hutto 2014). We categorize income into three classes (low, medium, high) and turn regression problems into classification problems (Kochhar 2018). The accuracy of our final income inference model is 70.02%.

#### Religious Status

We assign each user a boolean value for whether he/she is religious based on the tweets and the description in the profile (Zhang et al. 2020).

#### Family Status

By applying regular expression search, we identify users who show evidence that they are either fathers or mothers (Zhang et al. 2020).

#### Political Affiliations

The political attribute is labelled based on whether this Twitter user followed the Twitter accounts of the top political leaders. The president elect (Joe Biden^12^) and the incumbent president (Donald Trump) are included in the analysis.^13^

#### Sentiment

In our study, we intend to infer the sentiment of personal pandemic experience and non-pandemic experience. First, we use keyword search methods to classify the three-month historical tweets into COVID-related and non-COVID-related. If a tweet does not contain any of the keywords: “corona”, “covid”, “covid19”, “coronavirus”, “chinese virus”, “china virus”, “wuhan virus”,”wfh”, “work from home”, “pandemic”, “epidemic”, “herd immunity”, “quarantine”, “lockdown”, “mortality”, “morbidity”, “social distancing”, “mask”, “social distance”, “respirator”, “state of emergency”, “ventilator”, “isolation”, “fatality”, “community spread”, “vaccine”, “vaccinated”, “vaccination”, “panic buying”, “hoard”, it is categorized as non-COVID-related. The example tweets are

~~~
“< *user* > I could not WAIT to take
my husband’s last name! It was SUCH a
great feeling to solidify our union
by taking his name. Also kinda cringe
at the whole \keep the maiden name on
social media” thing some girls do…I’m
more \leave-and-cleave” type.”
~~~

and

~~~
“*< user >* what a glorious day that
was.”
~~~

The remaining tweets are categorized as COVID-19-related. The example tweets are

~~~
“i am now the kind of person who does
30 minutes of meditation and yoga from
my peloton app before settling into bed
to read a few chapters of my book and
be fast asleep before 11pm. quarantine
changed me.”
~~~

and

~~~
“*< user >* Oooorr…I can wear a mask,
get on an airplane, in a confined
space, with NO social distancing,
with people from HUNDREDS of different
households, ALL going to different
destinations, and then take my mask
OFF to eat/drink once I’m in my seat
*< hashtag > < hashtag >*“
~~~

For each Twitter user, the tweets of the two categories are concatenated, respectively. Next, a normalized, weighted composite score is calculated to measure the sentiment of the tweet content using Vader (Gilbert and Hutto 2014). The score is between −1 (most extreme negative) and +1 (most extreme positive).

### M3 Opinion Mining

To capture the opinions expressed through text by Twitter users, we adopt a human-guided machine learning framework inspired by Sadilek et al. (2013). The text are classified into four categories: (1) pro-vaccine, (2) vaccine-hesitant, (3) anti-vaccine, and (4) irrelevant.

To collect as many related text as possible, both COVID-19 vaccine-related and vaccine-related search keywords are used. However, the tweets collected using the vaccine-related search keywords are not necessarily related to COVID-19 vaccines. For example, MMR vaccine-related or HPV vaccine-related tweets might be crawled as well. In addition, the data collection is carried out during the flu shot season, resulting in collecting many influenza shot-related tweets. We apply a keyword-based search in tweets to remove all the tweets containing MMR, autism, HPV, tuberculosis, tetanus, hepatitis B, flu shot or flu vaccine (4.0% removed).

Tweets might be retweeted for multiple times. We observe that there are 6,703 non-unique tweets in the initial batch of over 90,000 tweets. These non-unique tweets, combined with their retweets constitute 62.9% of all tweets. As a result, the tweets are divided into two groups - the unique-tweet group and the non-unique-tweet group. 430 non-unique tweets which have been retweeted for at least 20 times are included in the non-unique-tweet group. These tweets and their retweets constitute 41.5% of all tweets. The rest are included in the unique-tweet group. All the tweets of the non-unique-tweet group are manually annotated. However, only a subgroup of the unique-tweet group are manually annotated. The state-of-the-art transformer-based language model (Yang et al. 2019), trained with the subgroup, is used to make estimates of the rest of the unique-tweet group.

#### Annotation

Three researchers read a sample corpus of tweets, discuss and set the standard of the opinion categories. Table 4 describes the standard for each opinion category. We label each tweet as one of the categories as long as it matches one of the descriptions of that category. During the labelling, three researchers independently read the text and make a judgement. The label is determined based on the consensus votes. However, in the case each researcher votes differently, the senior researcher determines the label for this piece of text after discussing with the other two researchers.

#### Tweets preprocessing

We adopt a tweet preprocessing pipeline from Baziotis, Pelekis, and Doulkeridis (2017) which can transform the specific text often used in Twitter to special tokens. For example, if the original tweet is

~~~
“Scientists develop a COVID vaccine
that could trigger a 10-times stronger
immune response *< url >* “
~~~

After preprocessing, the tweet becomes

~~~
“scientists develop a *< allcaps >* covid
*< /allcaps >* vaccine that could trigger a
*< number >* - times stronger immune
response *< url >* “
~~~

#### Performance of the XLNet model

Table 5 summarizes the performance of the final four-class XLNet model *H*_1_ on the external validation set with 400 samples. The final accuracy is 0.63 and the Kappa score is 0.5, which indicates a good agreement.

### M4 Analysis Details

#### Topic modeling

To capture the main concerns of the Twitter users regarding potential COVID-19 vaccines, we apply the Latent dirichlet allocation (LDA) topic modelling (Blei, Ng, and Jordan 2003). 10 topics with the top 30 keywords of each topic are presented in Figure 6. The coherence score is 0.31.

#### Factor indicators

To label the factor indicators, we use keyword search methods. The keywords for the safety and effectiveness include “safe”, “effective”, and “efficacy”. The keywords for the politics include “administration”, “politics”, “politician”, “political” and the names of Donald Trump, Mike Pence, Joe Biden and Kamala Harris.

## Footnotes

1 https://www.pewresearch.org/science/2020/09/17/u-s-public-now-divided-over-whether-to-get-covid-19-vaccine/

2 The distribution of the four categories is balanced.

3 https://help.twitter.com/en/managing-your-account/about-twitter-verified-accounts

4 https://uk.reuters.com/article/health-coronavirus-vaccine-poll/poll-more-women-than-men-in-us-nervous-about-fast-rollout-of-covid-vaccine-and-thats-a-problem-idUKL1N2IP361

5 https://www.pewresearch.org/science/2020/09/17/u-s-public-now-divided-over-whether-to-get-covid-19-vaccine/

6 Method section has detailed approaches.

7 https://www.nytimes.com/2020/08/02/us/politics/coronavirus-vaccine.html

8 https://www.tweepy.org/

9 The capitalization of non-hastag keywords does not matter in the Tweepy query.

10 https://github.com/CSSEGISandData/COVID-19

11 https://www.faceplusplus.com/

12 Joe Biden was the presidential candidate when the data was collected.

13 Due to limitation of Twitter API, only about half of Donald Trump’s follower ID was crawled.

